# Ni Binti Ni Mama: Participatory, human-centred development of a targeted nurturing care model for teenage mothers and their children in Kakamega County, Kenya – a mixed-methods study

**DOI:** 10.64898/2026.11.17.26368359

**Authors:** Everlyne Nyanchera Morema, Morris Senghor Shisanya, Sally Kosgey Oronje, Gregory Sakwa

## Abstract

Teenage mothers and their children face multiple barriers to nurturing care, yet few models translate the Nurturing Care Framework’s ‘targeted support’ level into practice for this group in sub-Saharan Africa. We describe the participatory, human-centred development of Ni Binti Ni Mama (‘she is a girl, she is a mother’), a targeted nurturing care model for teenage mothers in Kakamega County, Kenya. In phase one, a needs assessment combined a structured questionnaire and observation checklist covering the five nurturing care components (57 teenage mothers of infants), in-depth interviews with five mothers and five key informant interviews with health workers. Component scores were compared across groups using Welch’s t-tests; qualitative data were analysed thematically. In phase two, a stakeholder workshop with the sub-county health management team and university experts generated and prioritised strategies; a draft community health volunteer training module was tried out with 21 volunteers. The mean overall nurturing care score was 69.1% (SD 10.3). Uptake of maternal and child health services was high (good health score 88.1%), whereas early learning scored lowest (40.4%): 28.1% of children had a picture book and 15.8% of mothers had read or looked at pictures with their child the previous day. Most mothers (75.4%) had skipped meals for lack of money. Responsive caregiving was lower among mothers reporting emotional stress (76.4% vs 86.0%; p=0.005). Interviews described economic dependence, absent fathers, family conflict, peer stigma and food insecurity. The resulting model, framed by Social Cognitive Theory, combines community health volunteer home visits, school and family sensitisation, teen mothers’ clubs and peer mentorship, lactation support for school re-entry, and linkage to youth-friendly services and economic support. Teenage mothers informed the model through their needs and preferences but did not participate in design decisions; their direct involvement and a feasibility evaluation are the next steps.

## Introduction

Teenage pregnancy and consequent birth are a global problem which is worse in low- and middle-income countries, including Kenya [1]. In Kenya, 14.9% of women aged 15–19 years have ever been pregnant, and the proportion in Kakamega County is 15.1% [2]. Adolescent girls who have early and unintended pregnancies face many social, health and financial barriers to continuing with formal education, and poor school enrolment, retention, transition and completion have been attributed to high teenage pregnancy rates in some counties [3]. Teenage pregnancy and motherhood also pose a major risk to their children [4]: offspring of adolescent mothers have been reported to have more cognitive deficits and poorer growth than those of adult women [5], reflecting challenges uniquely experienced by this group, including inadequate support and mentorship on child care and difficulty sustaining breastfeeding. WHO recommends exclusive breastfeeding for the first six months, followed by complementary feeding with continued breastfeeding up to two years of age or beyond [6–8], which is difficult to achieve when young mothers return to school.

The stigma faced by adolescent girls with early and unintended pregnancies from peers and parents may affect their psychological well-being [9,10], resulting in depression during and after pregnancy. Maternal depressive emotions in the pre- and postnatal periods have been associated with children’s sleep problems [11]. Early identification of adolescent mothers and children at risk, alongside primary prevention of teenage pregnancy, is therefore necessary [12,13].

Sustainable Development Goal 3 targets an end to preventable deaths of newborns and children under five years of age by 2030 [14]. In 2018, WHO, UNICEF and the World Bank Group launched the Nurturing Care Framework to help children survive and thrive [15]. The Framework outlines three levels of support for families: universal, targeted and indicated. Teenage mothers are among the populations identified for targeted support, which aims to reduce the damaging effects of stress and deprivation, strengthen caregivers’ capacity to cope and provide extra help, such as home visits by trained providers, participatory community groups and affordable day care [15].

However, there is limited evidence on how targeted support can be designed for teenage mothers in sub-Saharan Africa, and existing parenting interventions for adolescent mothers have largely been developed in high-income settings [13]. Participatory, human-centred design approaches, which build interventions around the needs and preferences of intended users, offer a way to tailor such support to local contexts [16].

This study aimed to develop a targeted nurturing care model (Ni Binti Ni Mama) for teenage mothers and their children in Kakamega County, Kenya. The objectives were to: (i) assess the nurturing care practices, needs, challenges and preferences of teenage mothers; (ii) develop, with stakeholders, a model of targeted nurturing care informed by these findings; and (iii) develop a conceptual framework to guide evaluation of the model’s implementation.

## Materials and methods

### Study design

We used a sequential, mixed-methods, participatory human-centred design approach in two phases: (1) a needs assessment combining a cross-sectional survey and qualitative interviews; and (2) stakeholder-led model development. The study is reported following the GUIDED guidance for reporting intervention development studies [17] (S1 Checklist), and the COREQ checklist for the qualitative component [18] (S2 Checklist).

### Study setting

The study was conducted in the catchment area of Shibwe Sub-County Hospital, Idakho North Ward, Ikolomani Sub-County, Kakamega County, western Kenya. Ikolomani covers 143.6 km² in four wards (Idakho North, Central, South and East) and had a population of 111,743 at the 2019 census [19]. Ikolomani was purposively selected because it had the second-highest teenage pregnancy rate in the county (28%). During community entry at the sub-county headquarters, the team briefed the Sub-County Medical Officer of Health, Kakamega South, and filed the county research authorisation; the sub-county health management team recommended the Shibwe Sub-County Hospital catchment because it had the highest number of teenage mothers in the sub-county and well-established CHV networks; this area was therefore selected purposively, a change from the protocol, which had planned to sample all 28 community units in the sub-county. Participants were drawn from the Shibwe 1 and Shibwe 2 community units, which are linked to the hospital. Survey data were collected between 9 and 18 March 2023 and in-depth interviews between 5 and 16 April 2023.

### Phase 1: Needs assessment

#### Participants and recruitment

Teenage mothers were mapped with the help of community health volunteers (CHVs) and community health extension workers, and identified through CHVs and through chain referral by other teenage mothers in the two community units. Mothers aged 19 years or younger with a child under one year were eligible. The protocol specified a sample of 236, calculated with Fisher’s formula (n = Z²pq/d²) using Z = 1.96, an expected proportion of 19% (the national teenage pregnancy rate used at protocol development) and a margin of error of 0.05. Within the catchment and the data collection period, 68 survey interviews were completed. Four records submitted before the research licence was issued (test entries), one duplicate record and six mothers whose child was aged 12 months or older were excluded, leaving 57 mothers for analysis.

#### Quantitative data collection

A mobile-based structured tool, combining an observation checklist and an interviewer-administered questionnaire (S2 File), was administered on KoboCollect by trained research assistants; questions were asked in Kiswahili and responses translated back into English for recording. It was modified from standardised tools for measuring the components of the Nurturing Care Framework, drawing on suggested nurturing care indicators [20], taking into account the age of each respondent’s child. It covered socio-demographic characteristics; good health (10 items: family planning, HIV testing, PMTCT, antenatal care, skilled birth attendance, tetanus toxoid, child vaccination, counselling on child care, growth monitoring and care-seeking when the child was unwell); responsive caregiving (24 observed behaviours: 12 responsive, 4 non-responsive stimulation and 8 negative); opportunities for early learning (14 items on play materials and activities); safety and security (12 items on birth registration, food security, water treatment, housing, mosquito net use, handwashing, safe play spaces, financial support and protection from punishment and neglect); and adequate nutrition (early initiation and exclusive breastfeeding, continued breastfeeding for children aged 6 months or older, and maternal vitamin A). Five statements on maternal emotional stress (difficulty being affectionate, irritability, difficulty sleeping, nervousness and uncontrollable worry) were rated on a five-point agreement scale. Maternal weight and height and child weight, length and mid-upper arm circumference were measured using a tape measure, a MUAC tape and a digital weighing scale, and recorded to the nearest whole kilogram or centimetre. Child z-scores were calculated against the WHO 2006 Child Growth Standards and assessed against WHO data-quality criteria.

Items were scored 1 for the favourable response and 0 otherwise; non-responsive and negative caregiving behaviours, meal skipping and reduced meal size were reverse-coded, and agreement-scale items were scored 1 for agree or strongly agree. Each component score was the percentage of applicable items scored 1, and the overall score was the unweighted mean of the five component scores. The protocol’s three responsive caregiving sub-scores (responsive, non-responsive and negative behaviours, each as a percentage of items in that group) are also reported. Mothers were classified as reporting emotional stress if they agreed with at least one of the five statements. Anthropometric values outside plausible ranges were set to missing.

#### Qualitative data collection

In-depth interviews were conducted with five teenage mothers, and key informant interviews with five purposively selected informants: one community health extension worker, two lead CHVs, one sub-county community strategy focal person and one sub-county reproductive health nurse. Interviews with teenage mothers used a semi-structured guide on their experiences of motherhood, child care, support and challenges, and ideas for making care easier (S1 File), available in English, Kiswahili and the Luhya (Idakho) dialect. Interviews were conducted by the principal investigator with a note taker and were audio-recorded. During fieldwork, the qualitative component was expanded from the four interviews with teenage mothers planned in the protocol to five, and key informant interviews with community health workers and sub-county health staff were added to capture service-delivery perspectives. Mothers for in-depth interviews were randomly selected from survey respondents. Interviews were conducted mainly in Kiswahili at the mothers’ homesteads and lasted 18–28 minutes.

### Phase 2: Model development

Findings from phase 1 were presented in July 2023 at a dissemination and planning workshop, held at the county hospital as provided for in the approved protocol, attended by 15 participants: 11 members of the sub-county health management team, including the sub-county community strategy focal person, who represented CHVs, and four experts from the university. The workshop was used to: help stakeholders understand the rationale and findings of the needs assessment; generate proposals on the best course of action and on stakeholders’ roles; and gain support for programme design and implementation. The workshop opened with a presentation of the findings, after which participants’ views on the way forward were sought and decisions were reached by consensus. Proposed strategies were organised using Social Cognitive Theory [21] into a model and conceptual framework (Fig 1). A draft CHV training module on nurturing care was developed and tried out in a one-day workshop with 21 CHVs at Shibwe Sub-County Hospital.

**Fig 1.**
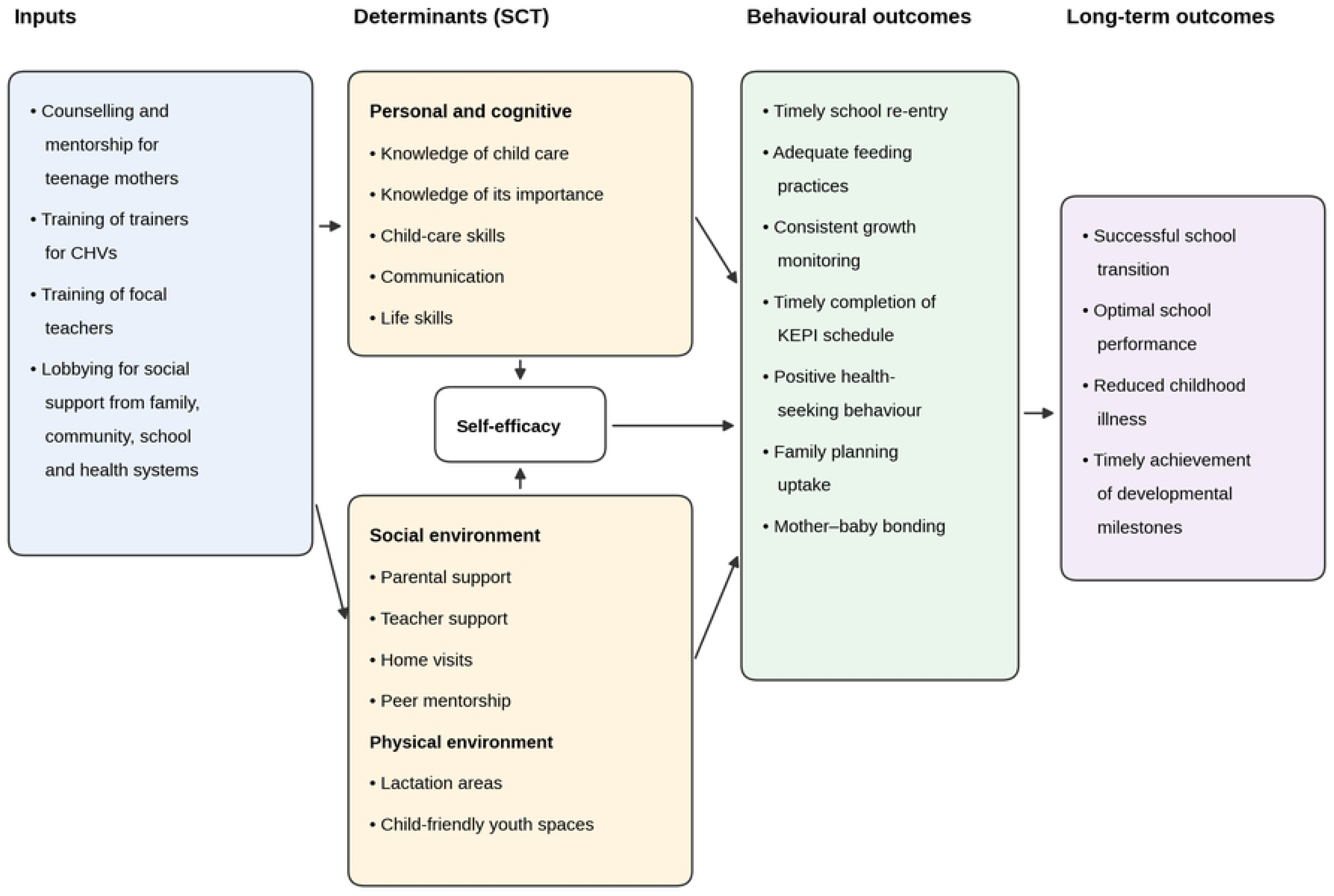
Ni Binti Ni Mama conceptual framework. Programme inputs act on teenage mothers’ personal and cognitive factors, self-efficacy and social and physical environment (Social Cognitive Theory) to produce behavioural outcomes for mother and child and longer-term outcomes; these outcomes provide the indicators for evaluating the model’s implementation. KEPI, Kenya Expanded Programme on Immunization; CHV, community health volunteer.

### Data analysis

Quantitative data were cleaned and analysed in Python 3.11 (pandas and SciPy); the de-identified dataset and analysis code are provided (S1 Data). Continuous scores are summarised as means and standard deviations, and categorical variables as frequencies and percentages. Mean scores were compared between groups using Welch’s t-test, with p<0.05 considered significant; given the sample size and number of comparisons, these analyses are exploratory. Internal consistency of the component scores was assessed with Cronbach’s alpha. Because scores were analysed as continuous variables, t-tests were used rather than the chi-square tests and logistic regression on categorised outcomes planned in the protocol. Interviews were transcribed verbatim, translated into English and analysed thematically. Quantitative and qualitative findings were integrated by triangulating them against the five nurturing care components, and a joint display (Table 4) maps identified needs to model components.

### Ethics statement

Ethical approval was obtained from the Masinde Muliro University of Science and Technology Institutional Scientific and Ethics Review Committee (MMUST-ISERC; approval no. MMUST/IERC/110/2023). A research licence was issued by the National Commission for Science, Technology and Innovation (NACOSTI/P/23/23603), and county authorisation by the County Government of Kakamega Ministry of Health Services (Ref. CGH/KAK/ERC/VOL.1/195). Participants under 18 years gave assent, and written consent was obtained from a parent or guardian; participants aged 18–19 years gave their own written informed consent. The information sheet was read aloud to participants who could not read, who indicated consent with a thumbprint. Key informants gave written informed consent. Participation was voluntary, participants could withdraw at any time, data were anonymised using unique codes, and electronic data were stored in password-protected, encrypted files. Verbal consent was obtained for audio recording, and referral arrangements were in place for participants who experienced distress.

## Results

### Participant characteristics

The 57 mothers had a mean age of 18.2 years (SD 1.1; range 15–19), and their children a median age of 5 months (Table 1). Half (50.9%) had secondary education, but 80.7% had dropped out of or paused schooling; 86.0% disagreed that their financial status was good, and 75.4% had parental support. Most households drew water from a river (54.4%) or spring (29.8%).

**Table 1.** Characteristics of teenage mothers and their children (n=57). IQR, interquartile range. *Derived from the mother’s and child’s ages. †One implausible value excluded (n=56).

| **Characteristic** | **Category** | **n (%)** |
|---|---|---|
| Mother’s age (years) | Mean (SD); range | 18.2 (1.1); 15–19 |
|  | 15–17 | 13 (22.8) |
|  | 18–19 | 44 (77.2) |
| Age at birth of index child (years)* | Mean (SD) | 17.8 (1.2) |
|  | ≤15 | 6 (10.5) |
|  | 16–19 | 51 (89.5) |
| Child’s age (months) | Median (IQR) | 5 (2–7) |
| Child’s sex | Female | 21 (36.8) |
|  | Male | 36 (63.2) |
| Mother’s own birth order† | 1st–3rd | 37 (66.1) |
|  | 4th or later | 19 (33.9) |
| Level of education | Primary | 22 (38.6) |
|  | Secondary | 29 (50.9) |
|  | Tertiary | 6 (10.5) |
| Schooling status | Continued | 11 (19.3) |
|  | Paused | 16 (28.1) |
|  | Dropped out | 30 (52.6) |
| Parental support | Yes | 43 (75.4) |
|  | No | 14 (24.6) |
| Financial status is good | Agree | 8 (14.0) |
|  | Neutral or disagree | 49 (86.0) |
| Receives financial support for child care | Yes | 34 (59.6) |
|  | No | 23 (40.4) |
| Main water source | River | 31 (54.4) |
|  | Spring | 17 (29.8) |
|  | Well | 9 (15.8) |

### Nurturing care practices

The mean overall nurturing care score was 69.1% (SD 10.3). Good health scored highest and opportunities for early learning lowest (Table 2).

**Table 2.** Nurturing care scores among teenage mothers (n=57). For the responsive caregiving sub-scores, the mean is the percentage of behaviours in each group that were observed; lower is better for non-responsive and negative behaviours.

| **Component** | **Items** | **Mean (%)** | **SD** |
|---|---|---|---|
| Good health | 10 | 88.1 | 9.0 |
| Responsive caregiving (composite) | 24 | 78.2 | 12.2 |
| Responsive behaviours observed | 12 | 86.3 | 19.1 |
| Non-responsive stimulation observed | 4 | 65.8 | 32.9 |
| Negative behaviours observed | 8 | 11.8 | 25.5 |
| Opportunities for early learning | 14 | 40.4 | 30.1 |
| Safety and security | 12 | 62.9 | 16.6 |
| Adequate nutrition | 2–3 | 76.0 | 26.5 |
| Overall (mean of five components) | — | 69.1 | 10.3 |

#### Good health

Uptake of maternal and child health services was high: 96.5% of mothers had attended antenatal care, had skilled birth attendance and had received tetanus toxoid, 96.5% of children were up to date with vaccination, all had growth monitoring recorded, 94.7% of mothers took the child for care when unwell and 93.0% had been tested for HIV. Lower proportions had received a PMTCT intervention (63.2%), were using family planning (61.4%) or had been counselled on child care (82.5%). Access to healthcare and family planning emerged as a key theme in the qualitative data: mothers obtained contraceptives free at the hospital but described side effects of the injection and implant, including heavy bleeding, weakness, hunger, dizziness and weight loss (IDI-3, IDI-4, IDI-5).

#### Responsive caregiving

Twenty-four observed indicators assessed responsive behaviour, non-responsive stimulation and negative caregiver behaviour. The composite score was 78.2% (SD 12.2). Most responsive behaviours were observed in over 75% of mothers, but fewer responded to the child’s cue with an open-ended question (63.2%). Non-responsive stimulation was common, for example praising the child in a non-responsive way (89.5%) or directing the child to a different activity (47.4%), while each negative behaviour, such as scolding, shouting or handling the child roughly, was observed in 3.5–15.8% of mothers.

#### Opportunities for early learning

This was the lowest-scoring component (mean 40.4%, SD 30.1). Singing to the child and talking with the child during housework were each reported by 70.2% of mothers, but only 28.1% of children had a picture book, 26.3% had objects of different shapes and colours, 28.1% had a designated place for playthings and 22.8% had received a new plaything in the past week; 15.8% of mothers had read or looked at pictures with the child and 36.8% had told stories in the previous 24 hours. In interviews, mothers described days taken up by housework (IDI-2, IDI-5) and economic dependence on their parents:

> *‘Maisha ni ngumu tu. Ni nadepend kwa wazazi. Hata diaper. Baba yake alitoroka akarudi kwao Tanzania na simu yake ni mteja. Chakula ni ngumu kupata na kazi hakuna. Kwa saa hii niko tu.’ (Life is just hard. I depend on my parents, even for diapers. His father ran away back to his home in Tanzania and his phone is unreachable. Food is hard to get and there is no work. For now, I am just here.) (IDI-1)*

#### Safety and security

The mean score was 62.9% (SD 16.6). All mothers slept under a mosquito net with the child, 75.4% washed their hands with soap before handling the baby and 71.9% reported a safe space for the child to play, but only 52.6% treated drinking water or had a well-ventilated sleeping room, and 66.7% of children had a birth notification or certificate. Most mothers agreed that they protected their child from physical punishment (94.7%) and neglect (93.0%), and 59.6% received financial support for the child’s care, most often from the child’s father (23 of 34) or their own parents (13 of 34). Food insecurity was the largest gap: 75.4% had skipped a meal and 87.7% had reduced meal size because of lack of money, and 49.1% could not access milk for the baby: ‘Chakula ni ngumu kupata na kazi hakuna’ (Food is hard to find and there is no work) (IDI-1).

#### Adequate nutrition

The mean score was 76.0% (SD 26.5). Most mothers reported early initiation and exclusive breastfeeding (86.0%), and 26 of 27 mothers of children aged 6 months or older were continuing to breastfeed; 59.6% had received vitamin A. In interviews, mothers linked difficulty with breastfeeding to food shortages: ‘Lakini chakula wakati mwingine hakipatikani na nikiwa na njaa maziwa huwa inapotea’ (But food is sometimes not available and when I am hungry, milk is lost) (IDI-1); another said that when food runs short ‘unalala njaa tu’ (you just go to bed hungry) (IDI-3). One mother was unsure how long she should breastfeed exclusively: ‘Huwa ninamnyonyesha tu vile waliniambia kwa clinic ingawa sijui ni kwa muda gani’ (I just breastfeed him as they told me at the clinic, although I don’t know for how long) (IDI-4). Mothers’ mean BMI was 24.9 kg/m² (SD 5.6; n=54); 7.4% had a BMI below 18.5 and 40.7% of 25 or above. Child anthropometric data did not meet WHO data-quality criteria and are not reported: weights and lengths had been recorded to the nearest whole kilogram and centimetre, 10 of 52 length-for-age z-scores (19.2%) fell outside WHO plausibility limits, and the standard deviations of the weight-for-age, length-for-age and weight-for-length z-scores (1.9–2.2) were well above the expected range of about 0.8–1.2.

### Factors associated with nurturing care scores

No socio-demographic characteristic was significantly associated with the overall nurturing care score (Table 3). In exploratory component-level comparisons, responsive caregiving was lower among mothers who agreed with at least one emotional stress statement (n=46) than among those who did not (76.4% vs 86.0%; p=0.005), and among mothers aged 18–19 years than those aged 15–17 years (76.4% vs 84.3%; p=0.014). Overall, 80.7% of mothers agreed with at least one stress statement; the most common were being unable to stop or control worrying (66.7%) and feeling nervous or anxious (45.6%).

**Table 3.**
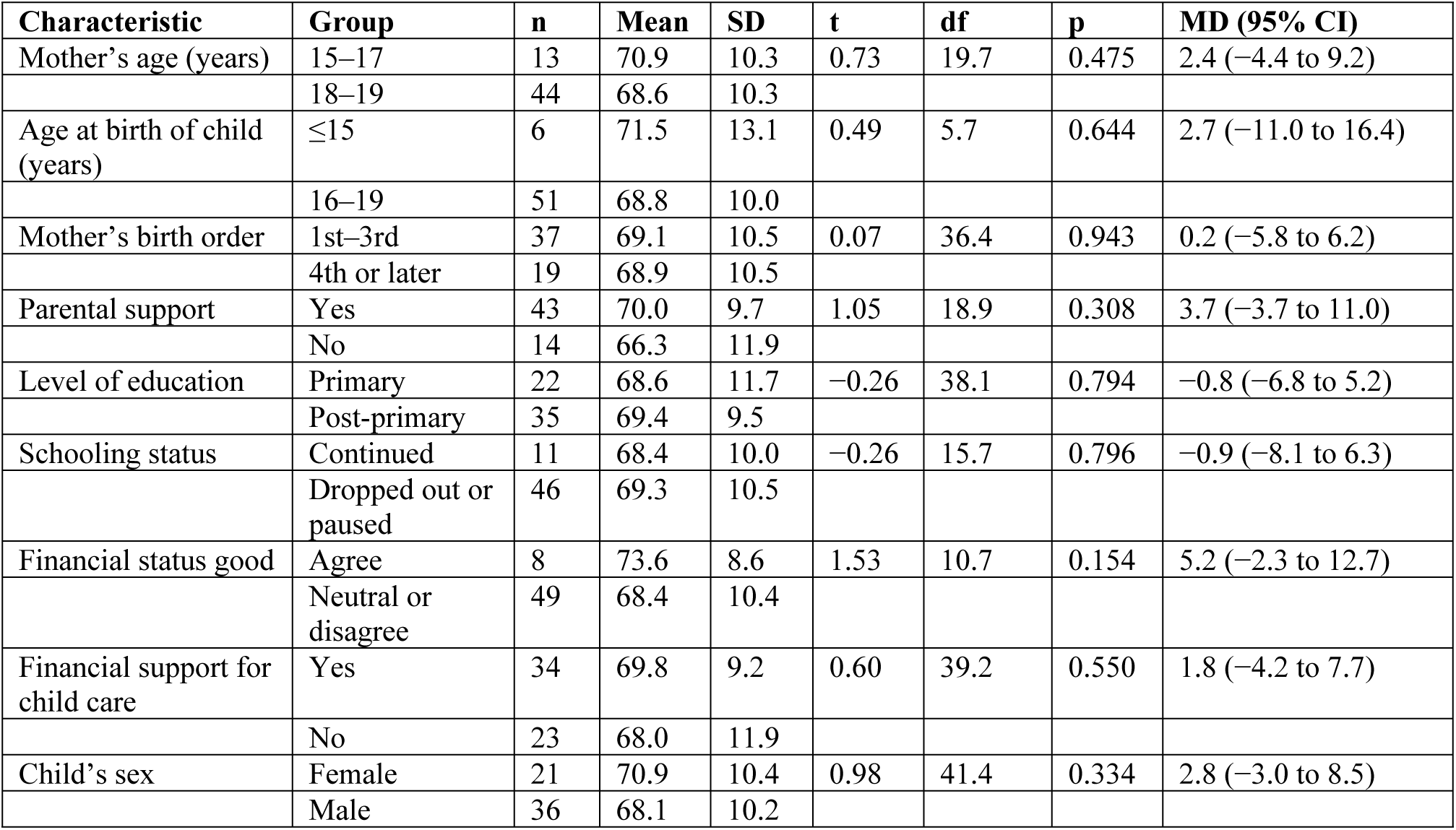
Mean overall nurturing care score (%) by characteristic (Welch’s t-test). MD, mean difference; CI, confidence interval.

| **Characteristic** | **Group** | **n** | **Mean** | **SD** | **t** | **df** | **p** | **MD (95% CI)** |
|---|---|---|---|---|---|---|---|---|
| Mother’s age (years) | 15–17 | 13 | 70.9 | 10.3 | 0.73 | 19.7 | 0.475 | 2.4 (−4.4 to 9.2) |
|  | 18–19 | 44 | 68.6 | 10.3 |  |  |  |  |
| Age at birth of child (years) | ≤15 | 6 | 71.5 | 13.1 | 0.49 | 5.7 | 0.644 | 2.7 (−11.0 to 16.4) |
|  | 16–19 | 51 | 68.8 | 10.0 |  |  |  |  |
| Mother’s birth order | 1st–3rd | 37 | 69.1 | 10.5 | 0.07 | 36.4 | 0.943 | 0.2 (−5.8 to 6.2) |
|  | 4th or later | 19 | 68.9 | 10.5 |  |  |  |  |
| Parental support | Yes | 43 | 70.0 | 9.7 | 1.05 | 18.9 | 0.308 | 3.7 (−3.7 to 11.0) |
|  | No | 14 | 66.3 | 11.9 |  |  |  |  |
| Level of education | Primary | 22 | 68.6 | 11.7 | −0.26 | 38.1 | 0.794 | −0.8 (−6.8 to 5.2) |
|  | Post-primary | 35 | 69.4 | 9.5 |  |  |  |  |
| Schooling status | Continued | 11 | 68.4 | 10.0 | −0.26 | 15.7 | 0.796 | −0.9 (−8.1 to 6.3) |
|  | Dropped out or paused | 46 | 69.3 | 10.5 |  |  |  |  |
| Financial status good | Agree | 8 | 73.6 | 8.6 | 1.53 | 10.7 | 0.154 | 5.2 (−2.3 to 12.7) |
|  | Neutral or disagree | 49 | 68.4 | 10.4 |  |  |  |  |
| Financial support for child care | Yes | 34 | 69.8 | 9.2 | 0.60 | 39.2 | 0.550 | 1.8 (−4.2 to 7.7) |
|  | No | 23 | 68.0 | 11.9 |  |  |  |  |
| Child’s sex | Female | 21 | 70.9 | 10.4 | 0.98 | 41.4 | 0.334 | 2.8 (−3.0 to 8.5) |
|  | Male | 36 | 68.1 | 10.2 |  |  |  |  |

### Qualitative findings

Six themes described the context of the mothers’ caregiving and informed the model. Family dynamics and struggles. Participants described disagreements with their parents, parental disapproval of the pregnancy and tension over childcare: ‘My parents were not happy that I was pregnant. They refused to let me get married but they were okay if the father and his people would come and take the child’ (IDI-5). ‘The mother always argues with me and even tells me to return the child to them. Sometimes she packs my clothes and tells me to go’ (IDI-5).

Social support and networks. Mothers mentioned parents, partners, grandmothers and sisters as sources of support: ‘Shosho yake na dada zangu wananisaidia kulea’ (His grandmother and my sisters help me raise him) (IDI-4); ‘Uhusiano wangu na wazazi wangu si mbaya. Wakanihimiza niolewe, wakaniambia nivumilie niweze kulea watoto wangu’ (My relationship with my parents is not bad. They encouraged me to get married and told me to persevere so that I can raise my children) (IDI-3). Most children’s fathers were absent or gave no support (IDI-1, IDI-2, IDI-4); one sent something only when called (IDI-5).

Gender roles and expectations. Traditional gender expectations shaped decisions about marriage and about returning the child to paternal relatives, as in the account of IDI-5 above. Mothers also saw contraception as the girl’s responsibility: ‘Wasichana huwa ndio wanatakikana kujichunga’ (Girls are the ones who are expected to take care of themselves) (IDI-2).

Youth and identity. Mothers described peer pressure and the emotional impact of others’ comments: ‘Mimi niliona nilikuwa mdogo. Na wenzangu walikuwa tu wananiongelea na nihisi vibaya’ (I felt I was young. And my peers were just talking about me and I felt bad) (IDI-5). Mothers aspired to further training or small businesses, such as hairdressing or growing vegetables (IDI-1, IDI-4).

Cultural and traditional influences. Cultural practices, such as the refusal to shave the child’s hair, caused unease among the mothers: ‘Nilikuwa vizuri lakini watu wa huko walikuwa wanakataa mtoto hata wakakataa kumnyoa nywele, lakini kijana mwenyewe akakubali akamnyoa’ (I was fine, but the people there [the father’s relatives] were rejecting the child and even refused to shave his hair, but the young man [the child’s father] himself agreed and shaved him) (IDI-5). Further probing revealed that shaving the child’s hair signifies acceptance by the paternal relatives, which matters because children are regarded as belonging to their fathers in this community.

Mental and emotional well-being. Mothers expressed discomfort, sadness and distress at negative comments from peers and from their own parents (IDI-5, above). Two had wished to end the pregnancy (IDI-1, IDI-3): ‘Nikiwa na mimba nilikuwa naogopa kutembea na hata kuenda shule. Nilifeel vibaya hata nikatamani nitoe’ (When I was pregnant, I was afraid to walk around or even go to school. I felt so bad that I wished to terminate it) (IDI-1).

### Stakeholder workshop and model development

Stakeholders emphasised the need to enhance parenting skills, foster emotional well-being and promote financial independence, noting that current government efforts focused on health and nutrition, with little attention to mental well-being, economic empowerment or parenting skills. The first intervention proposed was postpartum home visiting by CHVs, to whom teenage mothers would be referred after delivery for continuity of care, assessment and a support plan. This required strengthening CHVs’ capacity; a draft training module covering the Nurturing Care Framework, its five components and CHVs’ roles was therefore developed and tried out with 21 CHVs.

### The Ni Binti Ni Mama model

Ni Binti Ni Mama, loosely translated as ‘she is a girl, she is a mother’, aims to support teenage mothers to care effectively for their children while realising their own potential through school re-entry or economic activity. Social Cognitive Theory, which describes the influence of individual experiences, others’ actions and the environment on health behaviours, organised the strategies into a conceptual framework (Fig 1): building knowledge and self-efficacy, providing social support and role models, observational learning through peer mentors, reinforcement, and creating an enabling environment. Table 4 maps the needs identified in phase 1 to the components of the model.

**Table 4.** Joint display linking needs identified to components of the Ni Binti Ni Mama model.

| **Need identified (source)** | **Model component** | **Delivery agent** | **SCT construct** | **Nurturing care component** |
|---|---|---|---|---|
| Low early learning opportunities: few play materials and little reading or storytelling (survey); limited open-ended responses to the child (observation) | Postpartum CHV home visits with mentorship on nurturing care | CHVs | Knowledge, self-efficacy | Responsive caregiving; early learning |
| Family conflict, stigma (IDIs; KIIs) | Sensitisation of parents, teachers and community dialogues on teenage pregnancy and stigma | CHVs, focal teachers | Social support, environment | Safety and security |
| Emotional stress reported by 80.7% of mothers and linked to lower responsive caregiving (survey); distress and stigma (IDIs) | Teen mothers' clubs with monthly meetings; peer mentors | Peer mentors, CHVs | Observational learning, social support | Responsive caregiving |
| Food shortages affecting breast milk; uncertainty about duration of exclusive breastfeeding (IDIs); continuing breastfeeding on school re-entry (stakeholder workshop) | Lactation area in a model school; training on expressing, storing and transporting breast milk | Focal teachers, CHVs | Behavioural capability, environment | Adequate nutrition |
| Food insecurity (75.4% skipped meals) and economic dependence (survey; IDIs) | Cash transfers, then support for income-generating activities | County development fund | Reinforcement | Adequate nutrition; safety and security |
| High service uptake but lower PMTCT (63.2%) and family planning use (61.4%); contraceptive side effects (survey; IDIs; KIIs) | Teenage-friendly service hours; linkage to youth-friendly services; Social Health Authority cover for mother and child | Health facilities, SCHMT | Environment | Good health |

The expressed-breast-milk component proposes locally made insulated bags (crocheted and foil-lined) and frozen rice-filled packs to keep milk cool during transport and storage where refrigeration is unavailable. Expressed breast milk can be kept for up to 24 hours in an insulated cooler bag with frozen ice packs [22]; the performance of the locally made bags will be tested before roll-out.

## Discussion

This study describes the development of Ni Binti Ni Mama, a targeted nurturing care model for teenage mothers and their children, built from a mixed-methods needs assessment and stakeholder engagement in a rural Kenyan sub-county. Teenage mothers of infants made good use of maternal and child health services, but their children had few opportunities for early learning, and food insecurity and maternal emotional stress were common. These findings, together with the mothers’ accounts of economic dependence, absent fathers, family conflict and stigma, informed a model that extends beyond the health and nutrition focus of current services to include stimulation, psychosocial, educational and economic support.

Early learning was also a leading gap in an analysis of national survey data from Indonesia, in which most adolescent mothers provided incomplete nurturing care and a large group needed support particularly with nutrition and early learning opportunities [23]. In Kenya and Zambia, higher maternal stimulation was associated with better early child development, with language stimulation showing the largest effect [24]. The contrast between high service uptake and low early learning scores suggests that contact with health services has not yet translated into support for play, reading and storytelling, one of the five components of the Nurturing Care Framework [15]. This supports the model’s emphasis on CHV home visits that model responsive play and the use of low-cost, home-made play materials. Responsive caregiving was lower among mothers who reported emotional stress, consistent with the interview accounts of distress and stigma; this supports the inclusion of peer support and teen mothers’ clubs, and attention to maternal well-being during home visits. No socio-demographic characteristic was associated with the overall score, but the sample was too small to detect modest differences.

The model’s components align with the Nurturing Care Framework’s examples of targeted support – home visits for very young mothers, participatory groups and affordable childcare [15] – and complement mother–child programmes for adolescent mothers developed in high-income settings, such as the TeeMo intervention in Germany [13]. A distinctive element, proposed at the stakeholder workshop, is support for continued breastfeeding on school re-entry, which responds to Kenya’s school re-entry guidelines [3] and with the Health Act 2017 requirement for employers to provide lactation stations [25].

### Strengths and limitations

Strengths include the combination of quantitative and qualitative data from mothers and community health workers, observation of caregiving rather than self-report alone, grounding in an established framework and theory, and engagement of the sub-county health management team, which supports ownership and scalability. Limitations: the survey included 57 of the 236 mothers planned in the protocol, so it is underpowered and estimates are imprecise; data come from a single hospital catchment recommended by the sub-county health management team for its high number of teenage mothers and well-established CHV networks, and participants were recruited through CHVs and chain referral, so findings may not be representative; the survey was cross-sectional, comparisons were unadjusted and exploratory, and many were made; the good health and safety indices had low internal consistency (Cronbach’s alpha −0.11 and 0.53), reflecting near-universal service uptake, so they should be read as checklists rather than scales; maternal emotional stress was assessed with five unvalidated statements; child anthropometric measurements were recorded too imprecisely and with too many implausible values to estimate stunting, underweight or wasting, and maternal BMI is based on whole-unit measurements; self-reported practices may be subject to social desirability bias; teenage mothers informed the model but did not take part in the stakeholder workshop where design decisions were made; the qualitative sample was small (five mothers); and the model has not yet been delivered.

### Implications and next steps

The next phase will involve teenage mothers and their parents directly in refining the model, followed by a feasibility study assessing recruitment, acceptability, fidelity and preliminary outcomes, with baseline and endline surveys. The CHV training module and focal-teacher curriculum will undergo expert review before adoption.

## Conclusions

Ni Binti Ni Mama translates the Nurturing Care Framework’s targeted support level into a context-specific model for teenage mothers and their children, informed by their needs and by local stakeholders. It is ready for refinement with teenage mothers and for feasibility testing in Kakamega County and similar settings.

## Data Availability

All relevant quantitative data are within the manuscript and its Supporting Information files. The de-identified survey dataset, codebook and analysis code are provided as S1 Data. Interview transcripts are not publicly available because they contain information that could identify adolescent participants in a small community. De-identified transcript excerpts are available to qualified researchers on reasonable request to the Masinde Muliro University of Science and Technology Institutional Scientific and Ethics Review Committee.

## Acknowledgments

We thank the teenage mothers who shared their experiences; the community health volunteers of the Shibwe 1 and Shibwe 2 community units and the community health extension workers; the Kakamega South Sub-County Health Management Team; the County Government of Kakamega Department of Health; and our mentors, Dr Anthony Ajayi and Hleziwe Hara.

## Supporting information

S1 Checklist. GUIDED checklist.

S2 Checklist. COREQ checklist.

S1 File. In-depth interview guide for teenage mothers.

S2 File. Data collection tool.

S1 Data. De-identified survey dataset, codebook and analysis code.

